# IVF success rates in men with different causes of infertility: real world evidence from a population registry

**DOI:** 10.64898/2026.03.26.26349446

**Authors:** Oisin Fitzgerald, Georgina M Chambers, Clare Boothroyd, Robert McLachlan

## Abstract

**Background:** Male factor infertility is present in around 40% of couples utilising assisted reproductive technology (ART). However, it is unclear how specific causes of male infertility impact the chance of successful ART treatment, with most research either treating male infertility as single diagnostic group or being isolated smaller-scale studies focused on the treatment and outcomes of specific diagnoses.

**Objective:** To study the impact of eleven specific aetiologies of male infertility on the chance of a clinical pregnancy in couples with known causes of male or female infertility following their first ART cycle.

**Material and methods:** Population-based (initiated ART in Australia and New Zealand in 2020-2022) cohort study assessing the impact of eleven male infertility diagnoses (idiopathic, Klinefelter syndrome, Y chromosome microdeletions, testis damage from cancer, testis damage from other causes, gonadotropin deficiency, congenital absence of the vas deferens/cystic fibrosis (CBAVD), other obstruction disorder, erectile dysfunction, and ejaculatory disorder) on the chance of a clinical pregnancy following a couple’s first complete ART cycle (all fresh and frozen-thawed embryo transfers arising from one episode of ovarian stimulation). Adjusted risk ratios comparing a couples undergoing ART solely for treatment of male infertility with couples undergoing ART solely for treatment of tubal disease were calculated for the chance of a clinical pregnancy following a complete ART cycle and following an attempted fertilisation procedure.

**Results:** A total of 39,053 couples were included, with male infertility present in 42.7% of cases, and the only cause of infertility in just under half of these cases. In more than three-quarters of male infertility cases the cause of infertility was unknown (idiopathic) or undiagnosed. Most couples undertaking ART for treatment of male infertility can expect similar success rates to couples seeking treatment for good prognosis female infertility diagnoses. However, those with Klinefelter syndrome and Y chromosome microdeletions had a 59.5% (aRR: 40.5% [95% CI: 16.9%-64.1%]) and 28.9% (aRR: 71.1% [95% CI: 45.5%-96.7%]) lower chance of a clinical pregnancy *per* initiated stimulation cycle compared to those with tubal disease as the only source of infertility. However, there was no difference once sperm was retrieved compared to other diagnoses tending to require surgical sperm retrieval, use frozen oocytes and necessitating ICSI.

**Discussion and Conclusion:** In this population-based study most couples undergoing ART because of male infertility had similar success rates to those undergoing ART for treatment of female tubal disease, except for patients with Klinefelter syndrome and Y chromosome microdeletion who had approximately half and three-quarters the chance of a clinical pregnancy due to failed sperm retrieval/survival, but no difference (accounting for the use of surgical sperm, ICSI and potentially frozen oocytes) in outcomes once sperm were available. While these finding are reassuring for most men presenting to an ART clinic with male infertility, with more than three-quarters of male infertility cases reported as being idiopathic, there is an urgent need for greater research on the causes, diagnosis and implications of male infertility.

## Introduction

Male factor infertility is present in around 40% of couples utilising assisted reproductive technology (ART), procedures such as in vitro fertilisation (IVF) and intracytoplasmic sperm injection (ICSI), that involve the in vitro handling of both human oocytes and sperm or of embryos (Newman et al., 2023; Zegers-Hochschild et al., 2017). Indeed, the most common variation to ART treatment, intracytoplasmic sperm injection (ICSI) was developed in the early 1990s to overcome severe male infertility. Along with surgical sperm extraction techniques, ICSI combined with surgical sperm retrieval has enabled previously functionally sterile men to have children (Silber et al., 1995) . However, despite these developments it is unclear how specific causes of male infertility impact the chance of successful ART treatment, with most research either treating male infertility as single diagnostic group or being isolated smaller-scale studies focused on the treatment and outcomes of specific diagnoses.

The largest source of evidence for case-mix adjusted estimates of the impact of male infertility on ART outcomes (typically a clinical pregnancy or a livebirth) have been ART clinical prediction models, which have treated male infertility as single condition. Within this literature male infertility is associated with a similar livebirth rate compared to common female infertility diagnoses such as tubal disease and endometriosis (Dhillon et al., 2016; Luke et al., 2014; McLernon et al., 2016; Nelson & Lawlor, 2011; Vaegter et al., 2017). However, treating male infertility as a single condition may mask a heterogenous effect.

When comparing specific male infertility diagnoses it is important to consider both the cause and presentation, as similar presentations (e.g. no sperm in the ejaculate) can arise from dramatically different causes with potential implications for treatment and prognosis (Mazzilli et al., 2022; Nieschlag et al., 2023). Male infertility can be grouped into pre-testicular, testicular and post-testicular causes, with further stratification possible by whether the cause is congenital or acquired (Mazzilli et al., 2022). Ultimately male infertility presents as low sperm concentration (oligospermia), no sperm in the ejaculate (azoospermia), low sperm motility (asthenospermia), disformed sperm (teratospermia), or varying combinations of these three defects.

Pre-testicular causes include hypogonadotropic hypogonadism (HH) (such as Kallman syndrome) resulting in oligospermia and/or asthenospermia (Silveira et al., 2002) and while hormone therapy is the first-line treatment (Fink et al., 2024), it is not always successful, and patients may require ART. A meta-analysis found a 46% pregnancy rate among 709 congenital HH patients (Gao et al., 2018), with smaller case studies suggesting successful outcomes for men with acquired HH (Milsom et al., 2012).

Testicular causes may arise from genetic disorders impairing sperm production or direct testicular damage. Genetic causes include Klinefelter (47,XXY) syndrome (Aksglæde & Juul, 2013) and Y chromosome microdeletions in which specific Y chromosome deletions impair or completely halt spermatogenesis (Foresta et al., 2001; Krausz & Riera-Escamilla, 2018). Previous small scale (*n*<20) research has suggested patients with the most common Y chromosome deletions (AZFc deletion (Rabinowitz et al., 2021)) have comparable ART outcomes to otherwise similar fertile men (Choi et al., 2004), but highlighted difficulties in sperm retrieval as a major limiter of success in Klinefelter syndrome patients (Ulug et al., 2003).

Non-genetic causes of testicular damage include cancer, cryptorchidism and vascular defects such as varicoceles, with the level of reported success from ART depending on the degree of damage and impact of prior treatment (Chung & Brock, 2011; Esteves et al., 2010; Haimov-Kochman et al., 2010; Maheshwari et al., 2022). A history of cryptorchidism is commonly represented in the male with infertility, and in cases of severe impact can be treated with surgical sperm removal and ICSI (Chung & Brock, 2011), with reported live birth rates of 20% per ICSI procedure (Haimov-Kochman et al., 2010). Where ART required for clinical varicocele live birth rates of 31-46% per ICSI procedure are reported with variation depending on whether the varicocele is treated prior to commencement of ART (Esteves et al., 2010; Maheshwari et al., 2022). A common source of testicular damage is cancer treatment, within these patients with continuing spermatogenesis high levels of success with ART are seen (a pregnancy rate of 57% per cycle in *n*=17 couples) (Rosenlund et al., 1998). In 23-50% of cases the cause of impaired spermatogenesis is unknown (Krausz, 2011; Nieschlag et al., 2023; Pandruvada et al., 2021; Pierik et al., 2000), with this group generally reporting high live birth rates per initiated egg retrieval cycle (25% to 43%) (Gunn & Bates, 2016).

In obstructive azoospermia (OA), typified by patients with a prior vasectomy, spermatogenesis functions normally resulting in comparable outcomes to otherwise similar fertile men following surgical sperm recovery (Amorocho Llanos et al., 2022; Wosnitzer & Goldstein, 2014). Similar favourable outcomes are found in patients with erectile dysfunction or ejaculatory disorders (arising for instance from spinal cord damage or psychosexual issues) (Denil et al., 1996; Reignier et al., 2018).

In summary, while there are several isolated small studies on how specific male infertility diagnoses impact ART treatment and outcomes, larger population-level studies are lacking. This study aims to investigate the association between specific diagnoses of male infertility and the chance of a clinical pregnancy in couples with known causes of male or female infertility following their first complete ART cycle.

## Material and methods

### Data

The data for this study comes from the Australia and New Zealand Assisted Reproduction Database (ANZARD), a Clinical Quality Registry comprising information on all assisted reproductive technology (ART) treatment cycles undertaken in Australian and New Zealand fertility clinics. This study uses ANZARD data from treatments conducted in 2020-2023. In 2020 the ANZARD data collection was updated to record 12 diagnostic categories for male infertility.

This analysis investigates the outcome of a male-female couple’s first ever complete ART cycle, undertaken to treat medical infertility. A complete ART cycle is defined as an ovarian stimulation, or series of ovarian stimulations in which all oocytes are frozen, and all resulting attempted fresh and frozen-thaw embryo transfers (Chambers et al., 2017). This definition considers oocyte banking (i.e. a series of oocyte freezing procedures followed by thaw and attempted fertilisation) or oocyte freezing followed by thawing and concurrent oocyte pick-up (OPU) as part of the same complete ART cycle. Cycles in which all oocytes or embryos are frozen (“freeze-all” cycles) with no thawing during the follow-up period are included (i.e. denominators of the pregnancy rates) so long as they are not indicated as occurring for fertility preservation purposes.

Patients with the following characteristics were included in the study:

- A male-female intending parent couple using their own sperm and oocytes.
- A diagnosis of male or female infertility
- Initiated their first stimulated ART cycle between the 1^st^ of January 2020 and the 31^st^ of December 2022, with an administrative censoring date of 31^st^ December 2023 giving a minimum follow-up time of one year post initial ovarian stimulation procedure

Patients and cycles with the following characteristics were excluded from the study:

- Undertaking ART for non-medical purposes (e.g. elective or medical oocyte freezing).
- Use of donor sperm, oocytes or a gestational carrier.
- The cause of couple’s infertility is unexplained, which differs from a diagnosis of idiopathic male infertility. Unexplained infertility refers to cases where following complete investigation the cause of the couple’s infertility remains unexplained with no evidence of either male (e.g. demonstrable semen quality defect) or female factors (e.g. endometriosis, low ovarian reserve. In contrast, a diagnosis of idiopathic male infertility is made in the presence of reduced sperm quality or other male reproductive defects.
- Records for patients with incompatibility between the recorded sperm retrieval procedure and male infertility diagnosis.
- Records for patients with a male infertility diagnosis of “genetic: other aneuploidies/single gene” causing impaired spermatogenesis (rationale explained below).

The impact of these inclusion/exclusion criteria are illustrated in Supplementary Figure 1, with 27.1% of patients otherwise meeting the inclusion criteria excluded due to the couple’s unexplained infertility, and 0.4% and 1.1% of the remaining cycles excluded due to either incompatibility between the recorded sperm retrieval procedure and male infertility diagnosis or presence of a male infertility diagnosis of “genetic: other aneuploidies/single gene”.

Those with a male infertility diagnosis of “genetic: other aneuploidies/single gene” were excluded due to evidence that this category has been misinterpreted by the ART clinics providing data to ANZARD.

Our primary outcome is a clinical pregnancy, defined as fulfilling at least one of: [1] ongoing at 20 weeks; [2] evidence by ultrasound of an intrauterine sac and/or fetal heart; [3] examination of products of conception reveal chorionic villi; or [4] a definite ectopic pregnancy that has been diagnosed laparoscopically or by ultrasound (Newman et al., 2023). While live birth is often the preferred outcome from ART outcome studies, it was not available for cycles undertaken in 2023 at the time of analysis. We include the following patient characteristics: male age, female age, all eleven male infertility diagnosis recorded in ANZARD excluding “genetic: other aneuploidies/single gene” (idiopathic incl. past/present cryptorchidism, Klinefelter syndrome, Y chromosome microdeletions, testis damage from cancer, testis damage from other causes, gonadotropin deficiency, congenital absence of the vas deferens/cystic fibrosis (CBAVD), other obstruction disorders, erectile dysfunction, ejaculatory disorders), female infertility diagnosis (tubal disease, endometriosis, other (includes fibroids, ovulation disorders, premature ovarian failure, advanced maternal age etc.)). Treatment data included: Number of oocytes collected at OPU, concentration of sperm sample used for fertilisation (10^6^/mL), site of sperm extraction, use of in vitro fertilization (IVF) or intracytoplasmic sperm injection (ICSI), use of pre-implantation genetic testing (PGT), reason for PGT (aneuploidy screening, single gene variation, chromosomal structural rearrangements, other), number of fresh and frozen-thaw embryo transfers occurring following the ovarian stimulation.

## Software

The analysis was carried out using R version 4.2 (R Core Team, 2022), using *data*.*table* for data storage and manipulation (Dowle & Srinivasan, 2023), *ggplot2* for data visualisation (Wickham, 2016) and statistical packages as noted below.

## Descriptive analysis

The demographic and treatment characteristics of the analysis cohort are described using counts (%) for categorical variables, and medians [25^th^ percentile (Q1); 75^th^ percentile (Q3)] for continuous variables stratified by whether a live birth occurred. We characterise the distributions of male age and sperm concentration (10^6^/mL) using density plots. For each male diagnosis we illustrate the stage of treatment reached using a bar plot of the percentage of cycles that attempted oocyte fertilisation using IVF or ICSI, fertilised at least one oocyte (in the opinion of the treating embryologist), transferred at least one embryo (cleavage or blastocyst stage) and achieved a clinical pregnancy.

## Statistical models

### Missing data

Some of the variables in the dataset were impacted by missingness. In 9.1% of cycles information on previous pregnancies was missing. For these cycles this information was imputed using female age, male age, the infertility diagnosis and clinical pregnancy. In 2.1% of cases the source of sperm was not available. For these cases, it was imputed as the most frequent source of sperm based on the male infertility diagnosis. In 3.8% of cycles that reached attempted fertilisation using ejaculate sperm the sperm concentration was not recorded. For these cycles this information was imputed using male age, infertility diagnosis, fertilisation method, and fertilisation rate.

### Clinical pregnancy *per* complete cycle

We estimate a hierarchical generalised additive model (logistic link) for the chance of a clinical pregnancy *per* complete cycle using the *mgcv* R package (Wood, 2017). This model has covariates (fixed effects): female age at cycle start, male age at cycle start, female presence of infertility indicator variable and male infertility diagnosis along with intercept random effects terms for the ART clinic and country (Australia and New Zealand). Female and male age were modelled as non-linear (thin plate regression splines (Wood, 2003)). We present the effect of male infertility as an adjusted risk ratio (aRR) with a reference comparison of tubal disease. Tubal disease was chosen as our reference comparison as it has the highest live birth rate amongst female factor infertility diagnoses (Newman et al., 2023). Standard errors and 95% confidence intervals for the aRRs were calculated using the delta method using the *marginaleffects* R package (Arel-Bundock, 2024).

### Clinical pregnancy *per* attempted fertilisation

To model the chance of a clinical pregnancy following an attempted fertilisation procedure the same methodology was used as with the chance of a clinical pregnancy *per* complete cycle, albeit with this model estimating the direct rather than total effect of the male infertility diagnosis (Pearl, 2014), with the set of control variables extended to include: the number of oocytes collected at OPU, the proportion of frozen oocytes used, the sperm source (ejaculate, testicular or epididymis) and the attempted fertilisation method (IVF or ICSI). Additionally, the idiopathic male infertility category was divided based on semen sperm concentration into idiopathic (normal sperm count): ≥15×10^6^/mL, idiopathic (mild oligospermia): 10-15×10^6^/mL, idiopathic (moderate oligospermia): 5-10 ×10^6^/mL, idiopathic (severe oligospermia): <5×10^6^/mL (Choy & Amory, 2020; Cooper et al., 2010) and idiopathic (surgical): cases where sperm was surgically extracted from the testicles or epididymis. We note that sperm concentration is the only semen analysis parameter present in the source database and is only reported in cases where attempted fertilisation took place.

### Additional models

#### Treatment progression

For any male infertility diagnostic categories that had lower (at α=0.05) clinical pregnancy rates *per* complete ART cycle than couples undergoing treatment for tubal disease we further investigated at which stages(s) of ART treatment this difference arose. We modelled the probability of patients *transitioning* between the following stages:

- Cycle initiation → attempted fertilisation
- Attempted fertilisation → successful fertilisation
- Successful fertilisation → embryo transfer
- Embryo transfer → clinical pregnancy

Here cycle initiation refers to ovarian stimulation, attempted fertilisation can be using IVF or ICSI, successful fertilisation is the fertilisation by sperm (in the opinion of the treating embryologist) of at least one oocyte, and embryo transfer is the occurrence of at least one embryo transfer procedure within the complete ART cycle. Occurrence of each of these stages was represented as a binary outcome, and so the same methods and reference group (tubal disease) were used as the clinical pregnancy *per* complete cycle model. In addition to the variables controlled for in the clinical pregnancy *per* complete cycle model (i.e. ages and infertility) we added the following (confounders or balancing) variables to the models once the information they contained was in the past of the outcome being modelled: number of oocytes retrieved at OPU, number of oocytes fertilisation, source of sperm, use of IVF or ICSI and transfer of embryo screened with PGT. For instance, no new variables were added to the “cycle initiation → attempted fertilisation” model whereas number of oocytes retrieved at OPU was added to the “attempted fertilisation → successful fertilisation” model.

#### Live birth rate *per* complete cycle

In the studied cohort clinical pregnancy was used as the outcome as live birth outcome data was unavailable for treatment that extended into 2023 (i.e. those utilising frozen oocytes or embryos). Nevertheless, we estimated the impact of male infertility diagnosis on live birth using the approach (model and covariates) as for clinical pregnancy *per* complete cycle. A live birth was defined as any complete expulsion or extraction from the mother of a baby showing evidence of life that is of 20 weeks or more gestation or 400 grams or more birthweight (Newman et al., 2023).

### Ethics

Approval for this project was obtained from the UNSW Sydney Human Research Ethics Committee (reference number: iRECS0859).

## Results

### Descriptive analysis

As shown in Table 1, there were 39,053 couples who commenced their first ovarian stimulation cycles between 2020 and January 2022 who had a diagnosis of male and/or female infertility. Male infertility was present in 42.7% of cases, and the sole cause in 19.5% of these couples. Amongst the 21.7% of cases where the cause of male infertility was known 45.5% had post-testicular causes resulting in obstructive azoospermia (OA) or oligospermia including a prior vasectomy and congenital absence of the vas deferens/cystic fibrosis (CBAVD) (Table 1 and Figure 1A). Just over one-third (35.1%) had a known testicular cause, nine out of ten (92.1%) of which were testicular damage due to cancer or other causes followed by 7.9% with known genetic causes (Klinefelter syndrome or Y chromosome microdeletions). In those with male infertility the percentage of cases that reached attempted fertilisation requiring testicular and epidydimal sperm retrieval were 10.6% and 2.8%.

**Table 1.** Characteristics of study cohort, patients with known male or female infertility undertaking assisted reproductive technology (ART), Australia and New Zealand 2020-2023.

|  |  | Treatment commencement | Attempted fertilisation | Clinical pregnancy |
| --- | --- | --- | --- | --- |
| Patient characteristics |  |  |  |  |
| **N** |  | 40197 | 35783 | 16641 |
| **Male age (years)** |  | 37 [33; 41] | 36 [33; 41] | 35 [32; 39] |
| **Female age (years)** |  | 35 [32; 39] | 35 [31.75; 38] | 33 [30; 36] |
| **Infertility diagnosis** | *Male only* | 8708 (21.7%) | 8004 (22.4%) | 4718 (28.4%) |
|  | *Combined male and female* | 9058 (22.5%) | 7963 (22.3%) | 3229 (19.4%) |
|  | *Combined female factors* | 3463 (8.6%) | 3048 (8.5%) | 1213 (7.3%) |
|  | *Endometriosis* | 3186 (7.9%) | 2975 (8.3%) | 1665 (10%) |
|  | *Other* | 13581 (33.8%) | 11794 (33%) | 4688 (28.2%) |
|  | *Tubal disease* | 2201 (5.5%) | 1999 (5.6%) | 1128 (6.8%) |
| **Male infertility diagnosis** | *Idiopathic* | 14041 (79.0%) | 12665 (79.3%) | 6346 (79.9%) |
|  | *Normal sperm count ($\geq 15 \times 10^6/\text{mL}$)* | - | 6809 (53.8%) | 3294 (51.9%) |
|  | *Mild oligospermia ($10\text{-}15 \times 10^6/\text{mL}$)* | - | 949 (7.5%) | 483 (7.6%) |
|  | *Moderate oligospermia ($5\text{-}10 \times 10^6/\text{mL}$)* | - | 1282 (12.1%) | 671 (10.6%) |
|  | *Severe oligospermia ($<5 \times 10^6/\text{mL}$)* | - | 3139 (24.8%) | 1640 (25.8%) |
|  | *Surgical sperm extraction* | - | 486 (3.8%) | 258 (4.1%) |
|  | *Klinefelter syndrome* | 36 (0.2%) | 17 (0.1%) | 8 (0.1%) |
|  | *Y deletion* | 48 (0.3%) | 42 (0.3%) | 19 (0.2%) |
|  | *Testis damage: Cancer treatment* | 419 (2.4%) | 382 (2.4%) | 192 (2.4%) |
|  | *Testis damage: Other* | 771 (4.3%) | 675 (4.2%) | 354 (4.5%) |
|  | *Gonadotrophin deficiency* | 192 (1.1%) | 168 (1.1%) | 83 (1%) |
|  | *CBAVD/Cystic fibrosis* | 133 (0.7%) | 119 (0.7%) | 65 (0.8%) |
|  | *Vasectomy* | 1352 (7.6%) | 1217 (7.6%) | 545 (6.9%) |
|  | *Obstructive disorder: other* | 214 (1.2%) | 178 (1.1%) | 102 (1.3%) |
|  | *Ejaculatory disorders* | 233 (1.3%) | 206 (1.3%) | 87 (1.1%) |
|  | *Erectile dysfunction* | 327 (1.8%) | 298 (1.9%) | 146 (1.8%) |
| **Parity** | *Parous* | 6211 (15.5%) | 5479 (15.3%) | 2341 (14.1%) |
|  | *Nulliparous* | 30318 (75.4%) | 26881 (75.1%) | 12195 (73.3%) |
|  | *Missing* | 3668 (9.1%) | 3423 (9.6%) | 2105 (12.6%) |
| Treatment characteristics |  |  |  |  |
| **Complete cycles that reached OPU** | *Yes* | 35783 (89.0%) | 35783 (100%) | 16641 (100%) |
|  | *No* | 4414 (11.0%) | - | - |
| **Number of oocytes retrieved at OPU** |  | 8 [4; 14] | 9 [5; 14] | 12 [8; 17] |
| Complete cycle reached attempted fertilisation | Yes | 32612 (88.6%) | 32612 (100%) | 14641 (100%) |
|  | No | 4196 (11.4%) | - | - |
| Source of sperm (male infertility cases only) | Ejaculate | - | 13678 (85.7%) | 6806 (85.6%) |
|  | Epididymis | - | 430 (2.7%) | 212 (2.7%) |
|  | Testicular | - | 1527 (9.6%) | 720 (9.1%) |
|  | Missing | - | 332 (2.1%) | 209 (2.6%) |
| Sperm concentration (millions per millimetre) |  |  |  |  |
| Attempted fertilisation method | ICSI | - | 19506 (54.5%) | 9267 (55.7%) |
|  | IVF | - | 16277 (45.5%) | 7374 (44.3%) |
| Number of oocytes that underwent IVF or ICSI |  | 7 [3; 12] | 8 [5; 13] | 11 [7; 16] |
| Number of oocytes fertilised |  | 4 [2; 8] | 5 [3; 9] | 7 [5; 11] |
| Number of embryo transfer procedures | Zero | 10915 (27.2%) | 6513 (18.2%) | - |
|  | One or more | 20484 (51%) | 20476 (57.2%) | 12040 (72.4%) |
|  | Two or more | 8798 (21.9%) | 8794 (24.6%) | 4601 (27.6%) |
| Treatment outcomes |  |  |  |  |
| Clinical pregnancy | No | 23556 (58.6%) | 19142 (53.5%) | - |
|  | Yes | 16641 (41.4%) | 16641 (46.5%) | 16641 (100%) |

**Figure 1.**
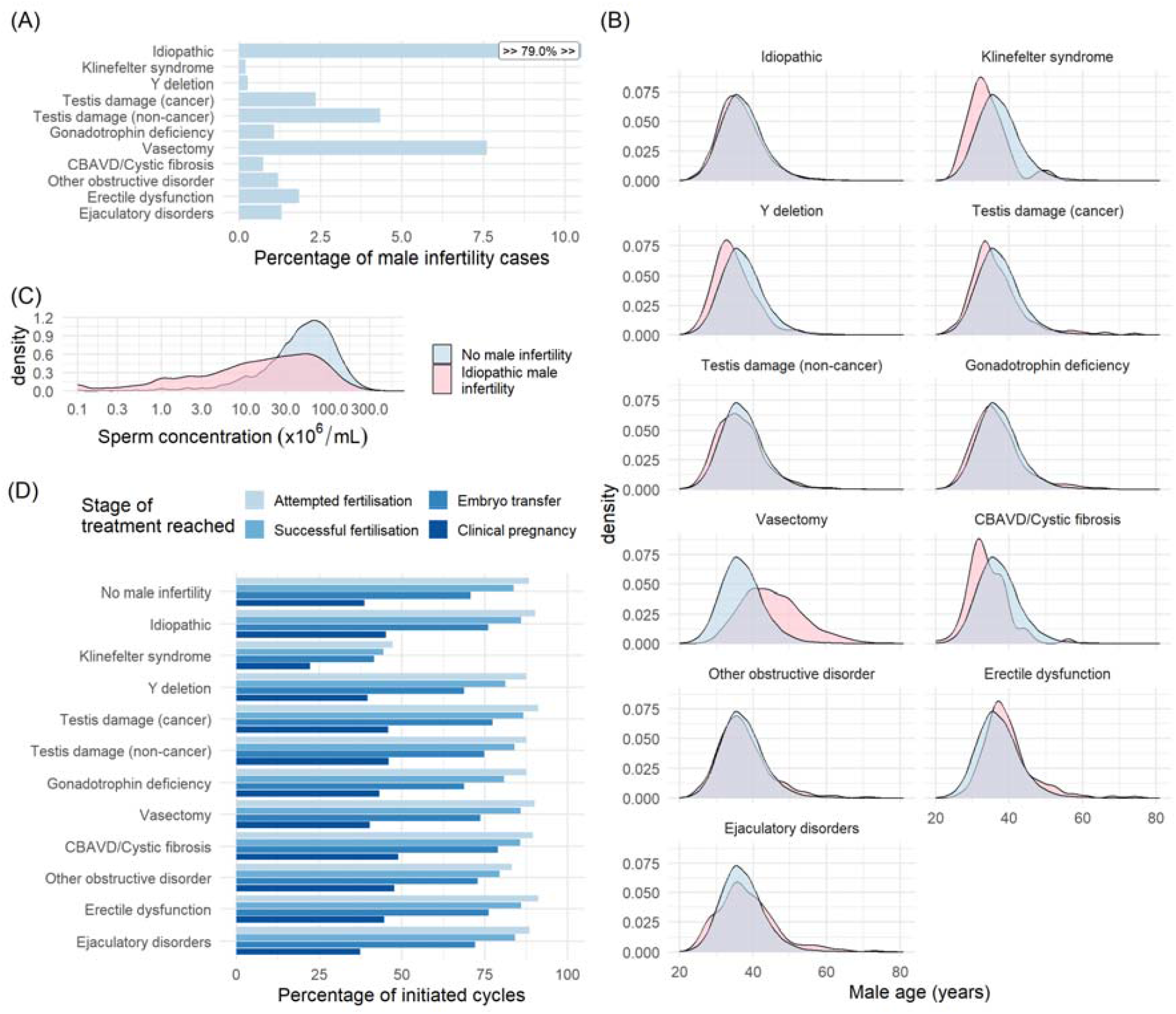
Characterisation of male infertility patients undertaking assisted reproductive technology (ART), Australia and New Zealand 2020-2023. A) Distribution of male infertility diagnoses. B) Age distribution by male infertility diagnosis (red), compared to those with no male infertility (blue). C) Sperm concentration (×10^6^/mL) (x-axis on log_10_ scale) in those with idiopathic male infertility compared to those with no male infertility. D) Percentage of initiated cycles that reached a particular stage of ART treatment by male infertility diagnosis. Attempted fertilisation could be with in vitro fertilisation (IVF) or intracytoplasmic sperm injection (ICSI). Successful fertilisation is the fertilisation of at least one embryo. Embryo transfer is the occurrence of at least one embryo transfer procedure. CBAVD: congenital bilateral absence of the vas deferens

The median [25^th^ percentile (Q1); 75^th^ percentile (Q3)] male age at treatment commencement was 37 [33; 41] years, and for female age was 35 [32; 39] years. The distribution of male age amongst infertile men was slightly younger than fertile men for most infertility diagnoses (Figure 1B), with the exceptions of men with a prior vasectomy (45 [39; 50] years), erectile dysfunction (38 [35; 42] years) and ejaculatory disorders (37 [34; 42] years) where the infertile men were typically older. Excluding these three diagnostic categories the median infertile male age was 36 [32; 40] years, compared to 37 [33; 41] years in fertile men.

Most (78.3%) cases of male fertility were due to impaired spermatogenesis of unknown cause (idiopathic male infertility), with this percentage even higher (82.1%) in couples with both male and female infertility. Men with idiopathic male infertility had median [IQR] ejaculate sperm concentrations of 20 [4; 42] ×10^6^/mL, significantly lower than the sperm concentrations of 51 [28; 84] ×10^6^/mL for non-infertile males undertaking ART (Figure 1C).

Oocyte freezing was far more common in Klinefelter syndrome and Y chromosome microdeletion patients, with 40.0% and 25.5% of these patients’ cryopreserving oocytes compared to the cohort average of 1.3%. Notably, despite the minimum one-year follow-up and indication that these were not fertility preservation cycles 55.6% and 64.3% of the Klinefelter syndrome and Y chromosome microdeletion couples who froze oocytes did not thaw them or attempt fertilisation using fresh oocytes. Here, fertility preservation is measured as an intention to store the oocytes for a year or more at the time of freeze.

The intermediate outcome and treatment characteristics of couples who achieved a clinical pregnancy were in line with existing literature (Table 1). They were more likely to: be younger with median [IQR] male and female ages of 35 [32; 39] years and 33 [30; 36] years compared to 37 [33; 41] years and 35 [32; 39] years in the complete cohort; have retrieved more oocytes at OPU (12 [8; 18] vs. 8 [4; 14]); fertilised more oocytes (7 [5; 11] vs 5 [5; 9]); and have had two or more embryos available for transfer (45.8% vs 29.0%).

Most male infertility diagnoses had broadly similar treatment progression statistics to fertile men (Table 1 and Figure 1D), with the exception of those with Klinefelter syndrome and Y chromosome microdeletion. Those with Klinefelter syndrome and Y chromosome microdeletion had a notably lower percentage of cycles reaching attempted fertilisation (53.5% and 70.9% compared to 86.5% in fertile men with an infertile partner) (Figure 1E) which translated into these groups achieving a lower clinical pregnancy rate *per* complete cycle than other patients (20.0% and 34.5% compared to 38.7%).

## Statistical models

Below we report the coefficients and parameters related to male infertility diagnoses, full coefficients and conditional spline effects for all models can be found in Supplementary Tables 1 to 12 and Supplementary Figures 1 to 6.

### Clinical pregnancy *per* complete cycle

The results of the statistical model for the chance of a clinical pregnancy following a couple’s first-ever ovarian stimulation cycle (complete cycle) were largely in line with unadjusted results in Table 1 and Figure 1D. Couples with Klinefelter syndrome (aRR: 40.5% [95% CI: 16.9%-64.1%]) or Y chromosome microdeletion (aRR: 71.1% [95% CI: 45.5%-96.7%]) as their only sources of infertility had a lower chance of a clinical pregnancy *per* complete cycle compared to couples with tubal disease and no male infertility (Figure 2a).

**Figure 2.**
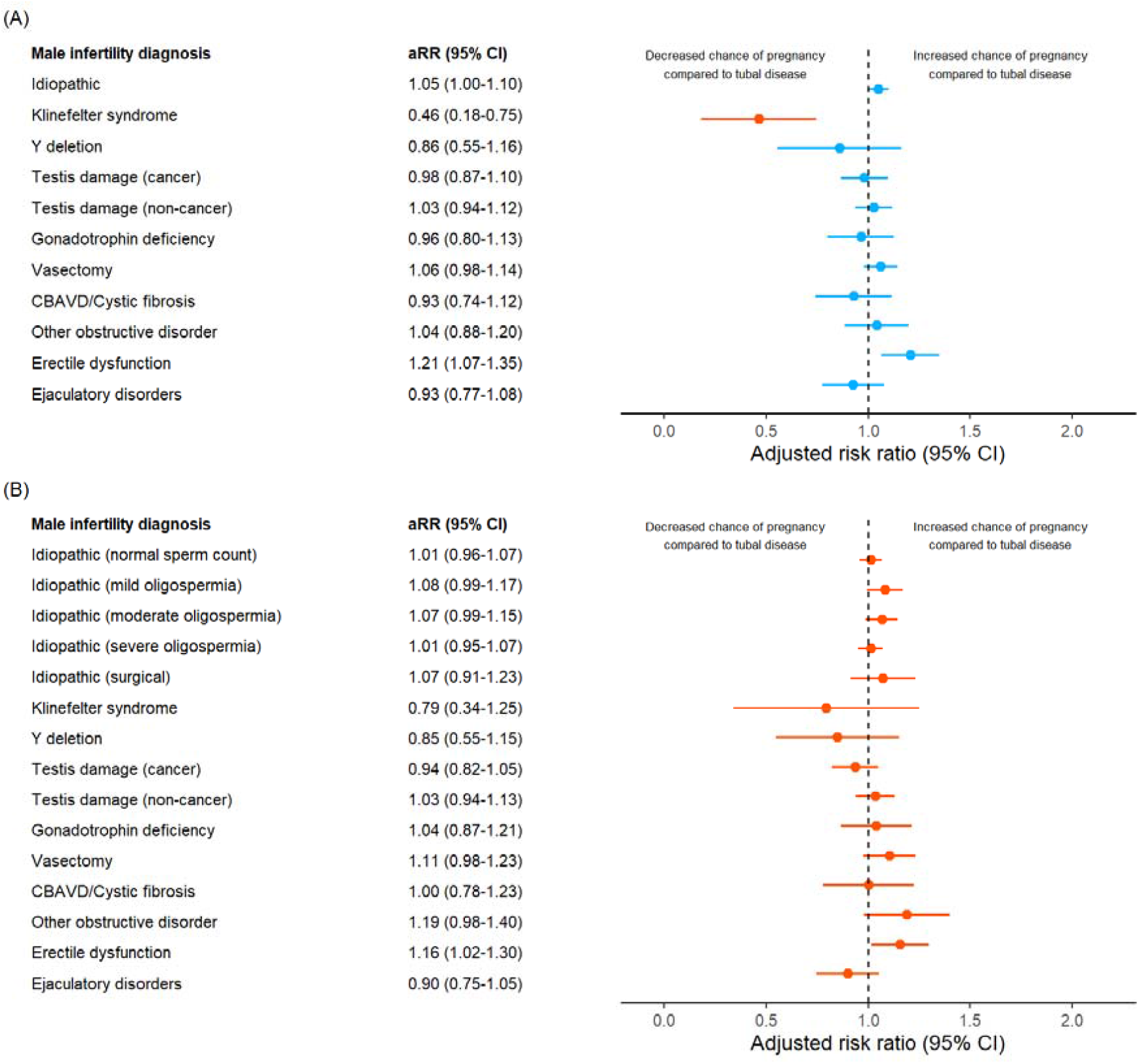
Results from the statistical analysis comparing couples undergoing ART solely for treatment of male infertility with couples undergoing ART solely for treatment of tubal disease. A) Adjusted risk ratio (aRR) for the relative effect of male infertility diagnosis compared to tubal disease on the chance of a clinical pregnancy following a complete (ovarian stimulation) assisted reproductive technology (ART) cycle. B) Adjusted risk ratio (aRR) for the relative effect of male infertility diagnosis compared to tubal disease on the chance of a clinical pregnancy following an attempted fertilisation procedure (using in vitro fertilisation or intracytoplasmic sperm injection) in a couples’ initial complete (ovarian stimulation) assisted reproductive technology (ART) cycle. Male and/or female infertility patients undertaking assisted reproductive technology (ART), Australia and New Zealand 2020-2023. CBAVD: congenital bilateral absence of the vas deferens

### Clinical pregnancy *per* attempted fertilisation

The results of the statistical model for the chance of a clinical pregnancy following a couple’s first-ever attempted fertilisation procedure are shown in Figure 2b with no male infertility diagnosis having a statistically significantly lower clinical pregnancy rate compared to the reference group.

### Additional models

#### Treatment progression

For Klinefelter syndrome and Y chromosome microdeletion patients we further examined at which stage of treatment their cycle was likely to fail by modelling their probability of progressing along the ART treatment pathway: *cycle initiation* → *attempted fertilisation* → *successful oocyte fertilisation* → *embryo transfer* → *clinical pregnancy*; comparing these estimates to patients with tubal disease. These results were also in line with the unadjusted results (Figure 1D), with Figure 3 showing that Klinefelter syndrome (aRR: 59.2% [95% CI: 42.7%-75.6%])) and Y chromosome microdeletion (aRR: 81.4% [95% CI: 68.1%-94.6%]) patients had a lower chance of progressing from cycle initiation to attempted fertilisation. All other treatment progression rates were similar to tubal disease patients.

**Figure 3.**
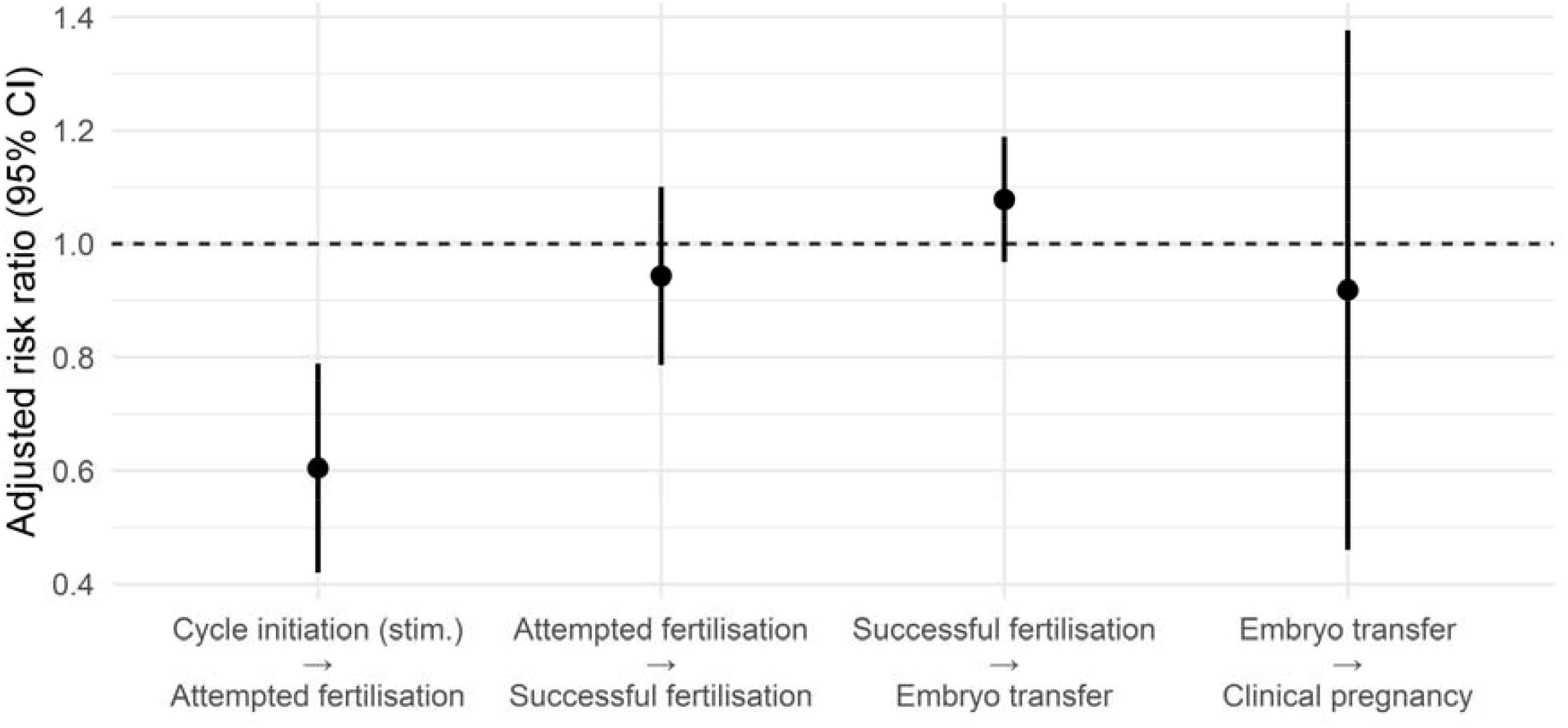
Adjusted risk ratio (aRR) for the relative effect of Klinefelter syndrome compared to tubal disease on the chance of treatment progression during a complete assisted reproductive technology (ART) cycle. Attempted fertilisation could be with in vitro fertilisation (IVF) or intracytoplasmic sperm injection (ICSI). Successful fertilisation is the fertilisation of at least one embryo. Embryo transfer is the occurrence of at least one embryo transfer procedure. Male and/or female infertility patients undertaking assisted reproductive technology (ART), Australia and New Zealand 2020-2023. Stim.: ovarian stimulation

#### Live birth *per* complete cycle

The findings for each male diagnosis in the statistical model for the chance of a live birth following a couple’s first-ever complete ART cycle (Supplementary Figure 2) were largely in line with the clinical pregnancy model (Figure 2), with Klinefelter syndrome (aRR: 44.2% [95% CI: 16.7%-71.7%]) patients having a lower chance of a live birth *per* complete cycle compared to couples with tubal disease and no male infertility.

## Discussion

The aim of this study was to investigate the effect of specific male infertility diagnoses on the chance of a clinical pregnancy in couples with known male or female infertility undertaking their first complete assisted reproductive technology (ART) treatment cycle. Our results suggest that most couples undertaking ART for treatment of male infertility can expect similar success rates to couples with seeking treatment for good prognosis female infertility diagnoses (tubal disease), once sufficient sperm have been obtained to commence treatment. However, those with Klinefelter syndrome and Y chromosome microdeletions had about half (aRR: 40.5% [95% CI: 16.9%-64.1%]) and three-quarters (aRR: 71.1% [95% CI: 45.5%-96.7%]) the chance of a clinical pregnancy *per* initiated complete (ovarian stimulation) cycle (Figure 2A). This was a result of a lower chance of progressing from cycle initiation (ovarian stimulation) to attempted fertilisation, presumably due to a combination of lower rates of successful fresh surgical sperm retrieval (SRR) and/or failed sperm thawing (Chen et al., 2020). We found no difference in clinical pregnancy rates once sperm was obtained for either group compared to other groups tending to require surgical sperm retrieval, use of frozen oocytes and necessitating ICSI (Figure 2B).

The current study adds direct comparisons of a wide range of male infertility diagnoses using population-level data, information lacking from prior research but largely supporting prior findings and implications. In line with population-level data targeted at informing patient counselling, such as the ART predictive modelling literature, the results suggest the majority of those with some form of male infertility can expect ART outcomes comparable to (or better than) couples’ undergoing treatment for tubal disease, in particular in cases where sperm has been successfully retrieved and attempted fertilisation undertaken.

This research highlights a need for ART registries to move away from only considering the female aspect of treatment (e.g. commencing data collection at ovarian stimulation) to recording male-related procedures which bear directly on ART success. These include surgical sperm retrieval (SSR), and other surgical or medical attempts to improve male fertility. Currently most ART registries do not record this valuable information despite male infertility being identified as an area in need of increased research (Kimmins et al., 2024). More than 13% of those with male infertility require SSR (Table 1). SSR for Klinefelter syndrome can fail nearly half the time (Kailash et al., 2021), with failure rates increasing with age, while for Y chromosome microdeletion patients success varies considerably depending on the chromosomal region affected (from low chances of sperm extraction for AZFa and/or AZFb deletions, to 45% failure rates or even use of ejaculate sperm for AZFc deletions) (Arshad et al., 2020). Incorporating this information into existing ART data registries would greatly inform clinical care and patient counselling for the 40% of couples affected by male infertility, and would go some way to redressing the disparity between male and female focused infertility research (Kimmins et al., 2024).

A notable finding of the current study was the high rate of idiopathic male infertility (78.3% of male infertility cases). Compared to fertile men this group had lower sperm concentration (Figure 1D), with no information available on other measures of sperm quality (e.g. motility or morphology). Nevertheless, their treatment outcomes were similar to patients being treated for tubal disease (Figure 2). While previous research has found or estimated lower rates of idiopathic male infertility rates, the much larger scale, and real-world nature, of the current research represents a key difference between the studies. Previous research (Pierik et al., 2000) has detailed how higher rates of specific male factor categorisation are reported following standardisation in areas such information technology (IT) system integration (e.g. between andrology and non-andrology services) and data collection. In contrast, in more routine practice it has been suggested that many male patients undergoing ART may not see an andrologist at all (Jequier, 2006). Indeed, in the current cohort the rates of idiopathic male infertility vary dramatically between ART clinics, from 57% to 97% in clinics treating over 200 patients a year suggesting wide ranging differences in clinical and diagnostic practice. This failure to develop and apply diagnostic tests for the cause of male infertility may be due to a tendency for infertile male patients to be treated primarily in an ART setting where male infertility risks becoming a cause (of a couple’s inability to conceive) rather than symptom of an underlying health condition, limiting detailed diagnostic investigations (Jequier, 2006; Krausz, 2011).

This study is not without limitations, some of which have been discussed above. The data source doesn’t enable determination of the rate of SRR failure, important information for those with conditions such as Klinefelter syndrome and Y chromosome microdeletions where this is the main point of treatment failure. Indeed, presumably the relatively large rate of oocyte cryopreservation for reasons other than fertility preservation without a subsequent thaw in Klinefelter syndrome and Y chromosome microdeletion patients was due to failed SSR (Lin et al., 2019). Further our data source doesn’t indicate if the sperm was fresh or frozen-thawed, although potentially this has little impact on outcomes (Mantravadi et al., 2025). Additionally, in this real-world data source most male infertility was recorded as idiopathic. This highlights the need for greater research into the genetic and acquired causes of impaired spermatogenesis, and a more multi-disciplinary view of male infertility (Jequier, 2006; Pierik et al., 2000). Indeed, given male infertility is a biomarker for somatic health and risk of future systemic illness, this systematic failure to investigate causes of male infertility limits opportunities to prevent further co-morbidity (Kimmins et al., 2024). This analysis primarily focused on the clinical pregnancy *per* complete cycle and attempted fertilisation. It is possible that as reported in prior research diagnoses such as Y chromosome microdeletions (Zhang et al., 2024) have impaired in vitro performance (e.g. fertilisation rate, embryo utilisation) which future diagnosis specific research will investigate. Further, live birth rather than pregnancy is generally considered the preferred outcome of studies on factors impacting ART outcomes. While the live birth outcome was not available for treatment cycles that persisted (using frozen oocytes or embryos) into 2023 the results from our live birth rate model were largely consistent with the clinical pregnancy findings (Supplementary Figure 2).

Strengths include the population-level nature of the study. The data source (ANZARD) collects information on all ART cycles that occur in Australia and New Zealand, enabling detailed determination of prevalence and outcomes of male infertility in those undertaking ART, and presentation of real-world evidence around the impact of male infertility diagnosis on ART treatment outcomes. To the best of our knowledge ANZARD is the only ART registry that enables clear differentiation of the cause and presentation of male infertility diagnoses, the importance of which is highlighted in the current findings – those with Klinefelter syndrome and obstructive disorders both present with azoospermia, but clearly have quite different prognoses. Further, by clarifying the exact stage of treatment in which Klinefelter syndrome is associated with reduced clinical pregnancy *per* complete cycle rates we provide quantitative evidence for use in counselling of patients and targets for further research.

### Conclusion

Most male infertility diagnoses have ART outcomes comparable to couples with a fertile male undergoing treatment for tubal disease. Patients with Klinefelter syndrome and Y chromosome microdeletions have reduced clinical pregnancy rates as a result of impaired sperm retrieval and/or survival, but no differences in outcome when sperm are obtained. While these finding are reassuring for most men presenting to an ART clinic with male infertility, there is an urgent need for greater research on the causes, diagnosis and implications of male infertility, with more than three-quarters of male infertility cases reported as being of unknown cause.

## Data Availability

The data that support the findings of this study are not publicly available due to their containing information that could compromise the privacy of participants.

## Author contributions

GC and RML conceptualised the study, acquired the funding and provided project supervision. OF curated the data, performed the formal analysis, designed the methodology and wrote the first draft of the manuscript. All authors reviewed and edited the manuscript.

---

**Supplementary Figure 1.**
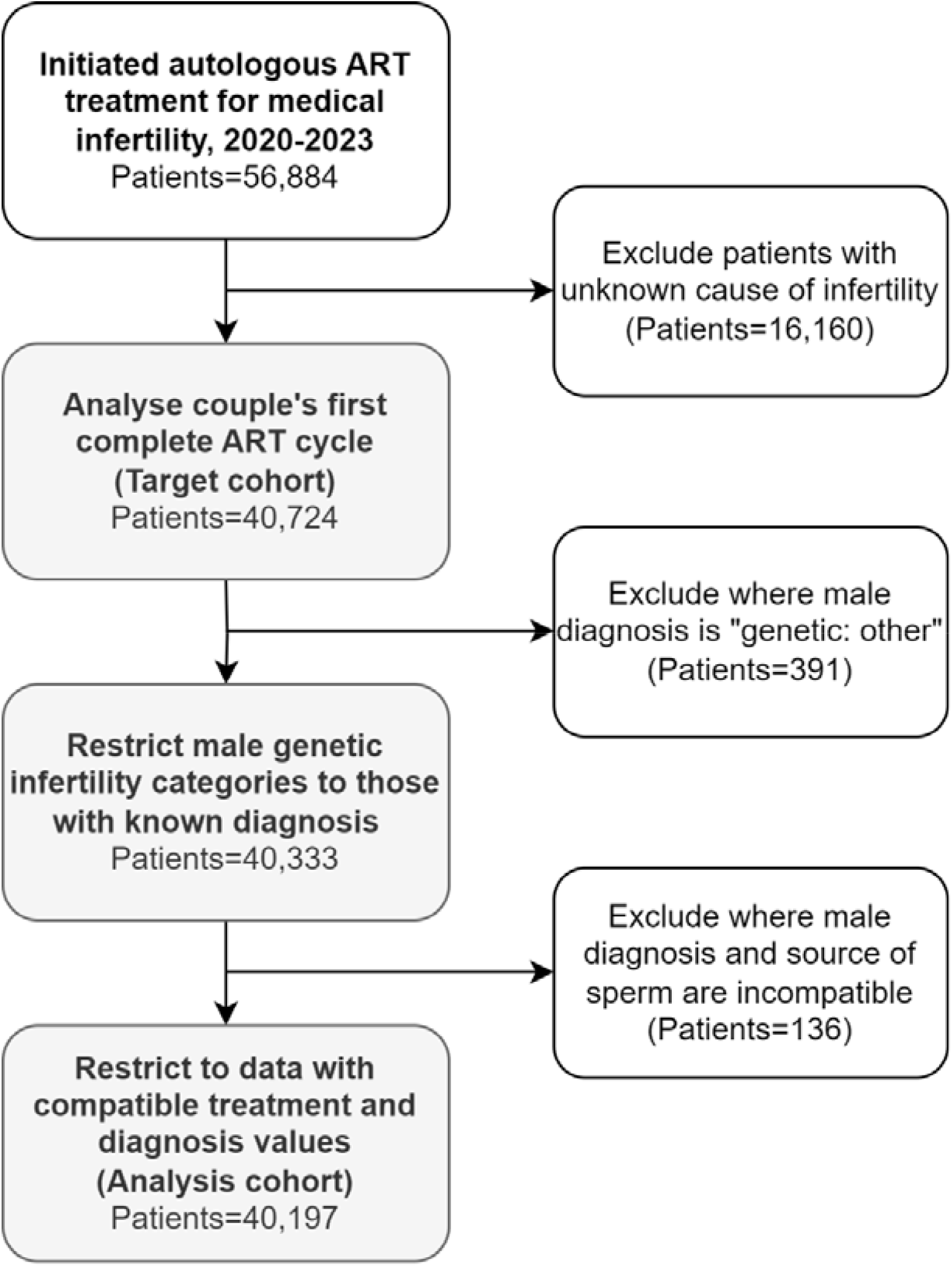
Impact of study inclusion and exclusion criteria and data quality exclusions on final analysis cohort. Male and/or female infertility patients undertaking assisted reproductive technology (ART), Australia and New Zealand 2020-2023.

**Supplementary Figure 2.**
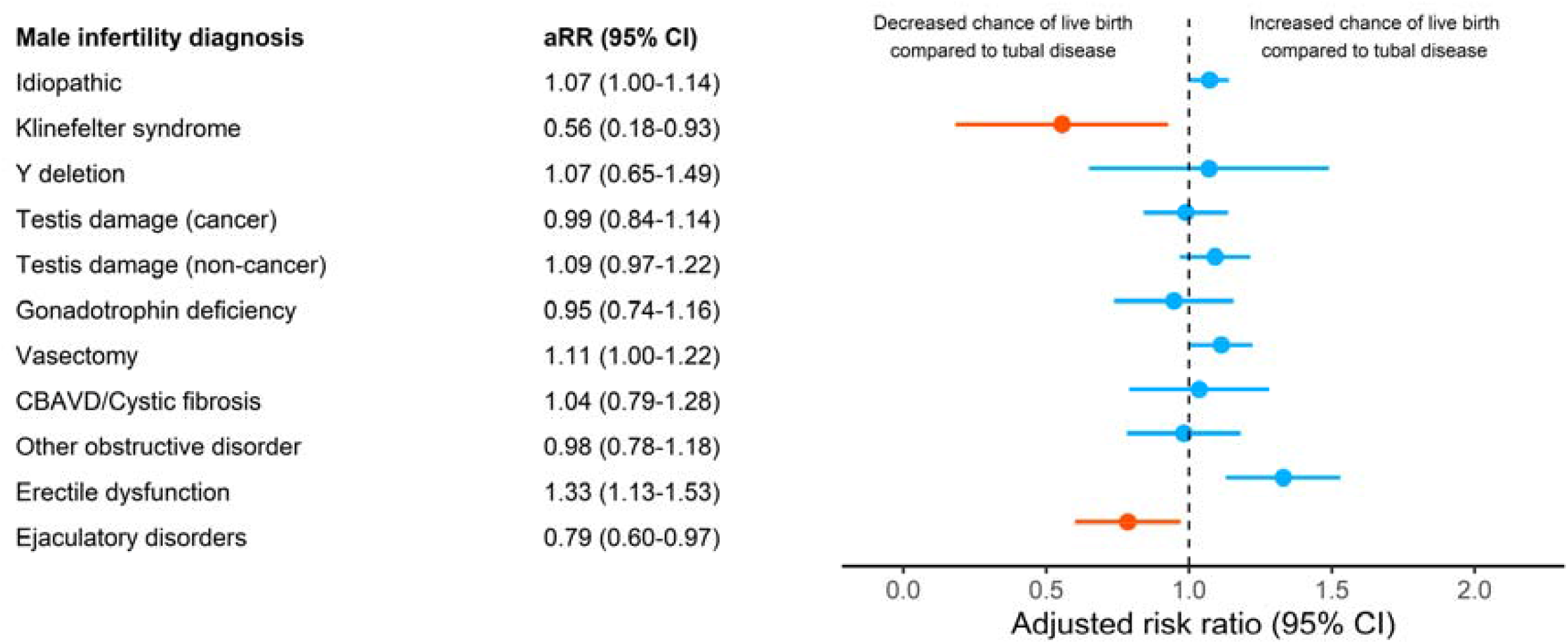
Adjusted risk ratio (aRR) for the relative effect of male infertility diagnosis compared to tubal disease on the chance of a live birth following a complete (ovarian stimulation) assisted reproductive technology (ART) cycle. Male and/or female infertility patients undertaking assisted reproductive technology (ART), Australia and New Zealand 2020-2022.

